# Improving Care by FAster risk-STratification through use of high sensitivity point-of-care troponin in patients presenting with possible acute coronary syndrome in the EmeRgency department (ICare-FASTER): a stepped-wedge cluster randomized trial

**DOI:** 10.64898/2026.04.21.26351433

**Authors:** Martin Than, John W Pickering, Laura R Joyce, Vanessa Buchan, Christopher M Florkowski, Nicholas L. Mills, Laura Hamill, Jason Prystowsky, Simon Harger, Mary Reed, Jared Bayless, Alexander Feberwee, Tamsin Attenburrow, Timothy Norman, Oliver Welfare, Tamika Heiden, Peter Kavsak, Allan S. Jaffe, Fred S. Apple, W. Frank Peacock, Louise Cullen, Sally Aldous, A. Mark Richards, Cameron Lacey, Richard Troughton, Christopher Frampton, Richard Body, Christian Müeller, Sarah Lord, Peter M George, Gerard Devlin

## Abstract

**BACKGROUND:** Point-of-care (POC) high-sensitivity cardiac troponin (hs-cTn) testing has the potential to expedite decision-making and reduce emergency department (ED) length of stay for patients presenting with possible myocardial infarction (MI) by ensuring that results are consistently available when looked for by clinicians. We assessed the real-life effectiveness and safety of implementing POC hs-cTn testing in the ED.

**METHODS:** We conducted a pragmatic, stepped-wedge cluster randomized trial. The control arm was usual care with an accelerated diagnostic pathway utilizing a single-sample rule-out step with a central laboratory hs-cTn assay. The intervention arm used the same pathway with a POC hs-cTnI. The primary effectiveness outcome was ED length of stay assessed using a generalized linear mixed model, and the safety outcome was 30-day MI or cardiac death.

**RESULTS:** Six sites participated with 59,980 ED presentations (44,747 individuals, 61±19 years, 49.5% female) from February 2023 to January 2025, in which 31,392 presentations were during the intervention arm. After adjustment for co-variates associated with length of stay, the intervention reduced length of stay by 13% (95% confidence intervals [CI], 9 to 16%. P<0.001), corresponding to a reduction of 47 minutes (95%CI, 33 to 61 minutes) from a mean length of stay in the control arm of 376 minutes. The 30-day MI or cardiac death rate was similar in the control and intervention arms (0.39% and 0.39% respectively, P=0.54).

**CONCLUSIONS:** Implementation of whole-blood hs-cTnI testing at the POC into an accelerated diagnostic pathway was safe and reduced length of stay in the ED compared with laboratory testing. (Australia New Zealand Clinical Trials Registry ACTRN12619001189112)

**Clinical Perspective:** *What is new?:*

- Point-of-care troponin assays with good precision have become available and received regulatory approval as high-sensitivity assays.
- A pragmatic stepped-wedge cluster randomized trial implementing the validated Siemens VTLi high-sensitivity point-of-care assay into 6 emergency departments investigating ∼60,000 patients, was conducted to ascertain if the rapid turnaround time reduced lengths of stay.
- Introduction of the VTLi high-sensitivity point-of-care assay into clinical pathways reduced emergency department lengths of stay by an average 13% (47 minutes) without compromising safety.

*What are the clinical implications?:*

- Early decision-making using a point-of-care high-sensitivity troponin assay within a structured clinical pathway was safe.
- Faster turnaround means troponin results are consistently available when clinicians first seek them. This can facilitate earlier decisions and reduce patient length of stay.

## INTRODUCTION

In 2022, 8.4 million presentations to emergency departments (EDs) in the USA were for chest pain and related symptoms of whom 378,000 (4.5%) had a myocardial infarction (MI).^1^ Rapid and safe assessment of patients with possible MI is necessary to expedite care and reduce ED length of stay. Key guidance bodies and colleges state reducing length of stay as a priority because of evidence associating ED crowding with adverse outcomes.^2–4^ This led to the design of accelerated diagnostic pathways (ADPs) combining cardiac troponin (cTn) measurements, an electrocardiogram, and clinical assessment that are now guideline-recommended globally.^5–8^ Single-sample rule-out (SSRO) strategies using high-sensitivity (hs-cTn) assays can safely facilitate rapid exclusion of MI in 30-50% of patients when carefully selected.

These strategies have reduced length of stay,^9^ but effectiveness has been limited by the time taken from blood-draw to result availability. When troponin results are not available for decision-making, at the completion of initial patient assessment, the clinician is likely to have proceeded to other tasks and be unable to action the troponin result promptly, even when it later becomes available. Point-of-care (POC) hs-cTn tests can be run using whole blood at or near the patient’s bedside and provide results within 8-20 minutes of blood draw. This can ensure that results are available when sought by the clinician and can expedite management decisions.

Improving Care by FAster risk-STratification through use of hs-cTn in patients presenting with possible acute coronary syndrome in the EmeRgency department (ICare-FASTER) was a pragmatic stepped-wedge cluster randomized trial. The aim was to evaluate the real-world effectiveness and safety of implementing hs-cTn POC testing in patients presenting to the ED with possible MI.

## METHODS

### DESIGN

ICare-FASTER was a pragmatic stepped-wedge cluster randomized trial. The protocol has been published and includes justification for the trial design.^10^ It describes the use of a sentinel (initial) site and run-in (transition) periods to guide, facilitate and embed the change in care. A large tertiary hospital (Christchurch) with experience of introducing ADPs was selected as the sentinel site (Fig 1A).^11–13^ Run-in periods were four months at the sentinel site and two months at other sites, during which the final training and live IT connectivity testing were performed and the intervention was made available and embedded into clinical practice. Six clusters each comprised of one site (hospital) began their control periods on 1 February 2023. The sentinel site had a 4-month control period. All other sites had longer control periods and were randomized to begin their run-in period at 1-month intervals. These periods were followed by intervention-periods of at least 5-months duration at each site. All sites finished on 31 January 2025 (Fig.1B). The control arm was the sum of all control-periods, and the intervention arm the sum of all intervention-periods.

**Figure 1.**
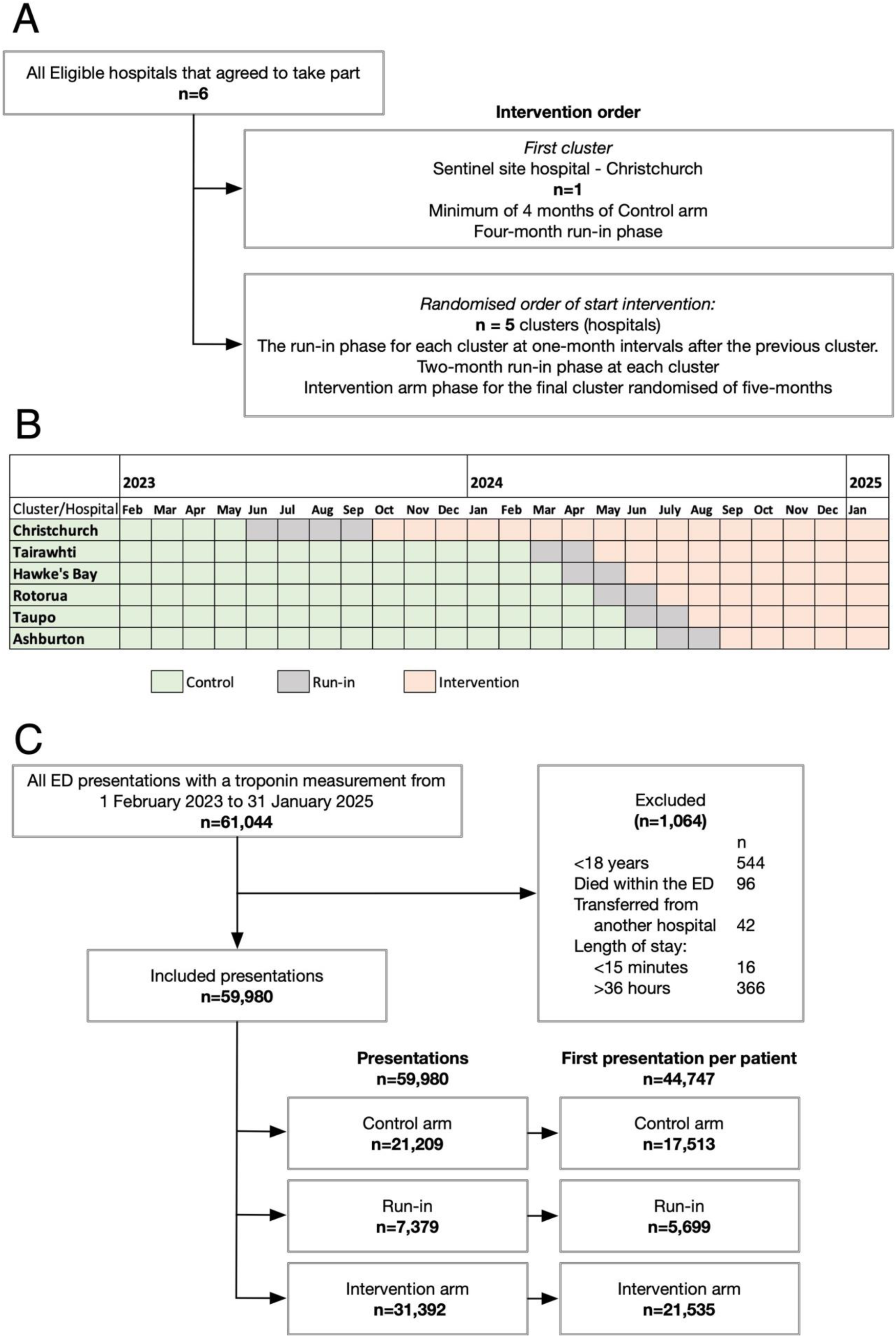
CONSORT diagram. (A) Cluster (Hospital) Randomisation. (B) Actual site control. Run-in, and Intervention periods, (C) All presentations analysed exclusion/inclusion.

### POPULATION

Sites were chosen to include small, medium and large hospitals, and to be representative of ethnic subgroups, particularly the Māori indigenous population. All sites already employed ADPs using a central laboratory hs-cTn assay and incorporating a single-sample rule-out step which, along with electrocardiogram and clinical assessment, identified a very-low-risk patients eligible for ED discharge. The time points for serial sampling were 0-hours and 2-hours. Participants were consecutive adults (≥18 years old) in whom a cTn test was performed for possible MI. The decision to perform cTn testing was made by the clinical staff responsible for care. Patients were excluded if they died within the ED or had been transferred from other hospitals.

### INTERVENTION

In the control arm, participants received usual care with laboratory hs-cTn assays to guide decision-making (see Supplementary table S1 for the assays used). In the intervention arm laboratory assays were replaced by a POC hs-cTnI assay as the default test. There were no other changes to the ADP steps. Fig.2 illustrates the ADP from one hospital, Supplemental Appendix Fig.S1.1 to S1.6 are the full pathways. Nurses performed the POC hs-cTnI testing as part of their usual clinical activities. The laboratory test could be used during the intervention period when a patient required further investigation prompting admission, or had a recent laboratory hs-cTn test so that further measurements could be compared with that baseline, or if the POC analyzer delivered an error message or was unavailable. When there was an error message, nurses usually repeated the test on the analyser with an option to send a sample to the laboratory for testing. In both instances, the existing blood sample was used (i.e. there was not an additional blood-draw).

**Figure 2.**
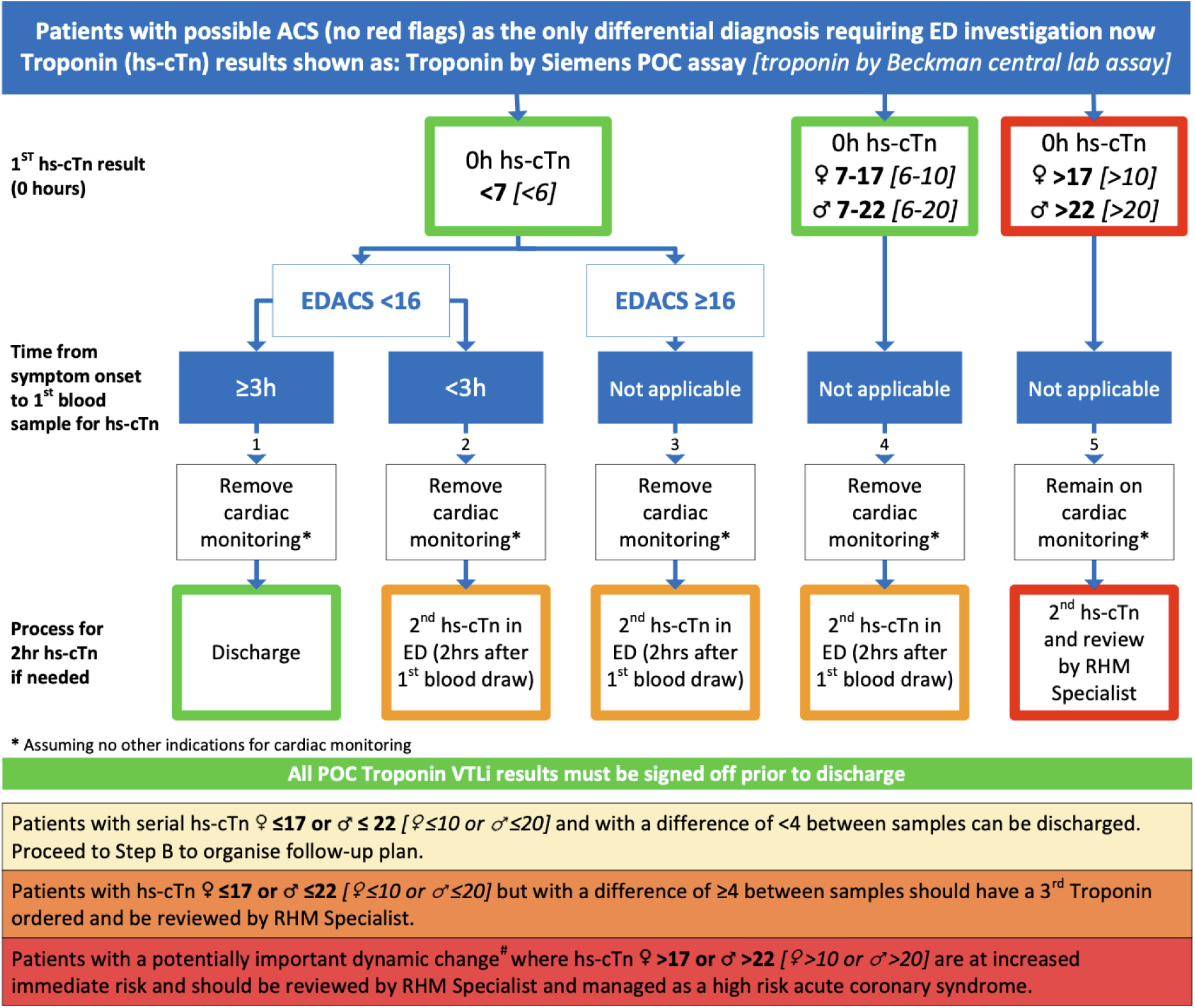
Accelerated Diagnostic Pathway schematic for one hospital. Numbers are ***POC assay*** *[Laboratory assay when used]*. Full pathways for all hospitals are in the Supplemental Appendix.

The interventional POC hs-cTn assay was measured in whole blood using the Siemens Healthineers Atellica VTLi hs-cTnI (Erlangen, Germany). The limit of detection is 1.6 ng/L, limit of quantitation 3.7 ng/L, female and male 99^th^ percentiles 18 ng/L and 27 ng/L and overall URL of 23 ng/L, and analytical turnaround time 8 minutes. Prior to implementation the assay’s analytical precision was verified,^14^ and a safe single-test low-risk threshold of 7 ng/L was determined for incorporation in ADPs.^15^ The control laboratory assays’ characteristics are given in Table S1.

### DATA COLLECTION

All cTn measurements were linked via the patient National Health Index, a unique identifier that links every person in New Zealand to all hospital events, deaths, and ICD10 codes. This was used to identify all admissions to any hospital, MI and death within 30-days of the ED presentation. This enabled complete follow-up of all patients remaining in New Zealand. In a previous stepped-wedge trial the use of National Health Index to identify these outcomes was found to have 98% agreement with adjudicated outcomes.^16^ The time of patient arrival and discharge or admission were routinely recorded within the electronic patient record.

### PRIMARY AND SECONDARY OUTCOMES

The primary outcome was the ED length of stay, measured in minutes, and defined as the difference between arrival time and the time of discharge home or hospital admission. All presentations (rather than only first presentations) were considered to reflect the real-life impact on ED occupancy.

Secondary effectiveness outcomes included performance in those discharged: (1) directly from ED, (2) after only a single cTn test, (3) after a single cTn test result below the low-risk threshold, and (4) after two or more cTn tests. Additionally, the change in odds for discharge within 1, 2, 3, 4, 5 and 6 hours from arrival was calculated.

Several sensitivity analyses were conducted, including where (i) only each patient’s first presentation during the study period were included, (ii) additional presentations from the run-in periods with the start of the intervention arm where the interventional assay was available for use were included, (iii) the non-randomized sentinel site was excluded, (iv) presentations in the intervention arm assessed with the laboratory assay were excluded, (v) the period where at least one site was allocated to a control period or run-in period and at least one site was in an intervention period, (vi) only those where the first blood draw was within one hour of ED arrival were included, (vii) exposure time was included in the model. Sub-groups included sex, ethnicity, and age-group.

Safety outcomes included MI or cardiac death following discharge within 30 days in unique patients discharged from the ED. MI was defined using ICD-10 codes, but each death was reviewed by two independent clinicians and adjudicated as either cardiac or non-cardiac death. Death was adjudicated to be cardiac if there was a clear cardiac cause or if there was no clear alternative cause. Secondary outcomes and sensitivity analyses were pre-specified (see the protocol^10^ and statistical analysis plan available at https://www.anzctr.org.au).

### STATISTICAL ANALYSIS

The primary outcome of ‘length of stay in the ED for all presentations’ was modelled using a generalized linear mixed model including: study arm, cluster (site), time from study start in days (secular trend), season, hospital shift, and time from presentation to first blood draw as covariates. We did not assume a linear change in secular trend or the same change at each site applied and so modelled it as a non-linear function using restricted cubic splines (more details are in the Statistical Analysis Plan). As length of stay was non-normally distributed it was log (base 2) transformed prior to analysis, and therefore the results are presented as geometric means and mean ratios. The geometric mean ratios and 95% confidence intervals (CIs) are presented as percentage changes and absolute differences for the intervention arm compared with the control arm. A sample size of 5,785 was sufficient to detect a ≥15 minute length of stay at an α = 0.05 with a power of 90%.^10^ A binomial generalized linear mixed model was used for discrete outcomes of discharge from ED prior to a specified length of stay. All analyses were undertaken using R v4.5, using the *lme4* package.^17,18^

Ethical approval was granted by the New Zealand Southern Health and Disability Ethics Committee, reference 21/STH/9. This paper used the CONSIDER statement^19^ to guide reporting on ethnicity and the CONSORT guidance for stepped-wedge cluster randomized studies^20^ and the StaRi guidance for implementation studies.^21^

## RESULTS

### SITES AND POPULATION

The six sites varied from a small regional hospital to a large tertiary hospital and were representative of the population (Table S2 and S3). Intervention periods ran for at least 5 months and up to 16 months at individual sites (Fig.1B). There were 59,980 presentations from 1 February 2023 to 31 January 2025 including the run-in phase in 44,747 individual patients (Tables S4, S5; Fig.1C). Over the two-year study period there was a 23.4% increase in total ED presentations across sites from 19,647 to 24,249 per month (Fig.S2). This was consistent with a national trend of increasing ED volumes (Fig.S3).

The mean age on first presentation was 61 years (range 18 to 103 years), and 49.5% were female (Table 1). There were 21,209 presentations in the control arm and 31,392 in the intervention arm (total 52,601) of whom 12,906 (60.9%) and 19,271 (61.3%) were discharged from the ED respectively. During intervention 17,065 (54.4%) were assessed by POC alone, 8,252 (26.3%) by laboratory test alone, and 6,075 (19.4%) by both tests (Fig.3). There were 37,610 POC tests across all blood-sample time points of which 34,394 (91.4%) gave valid results and were used in clinical decision making. There were 3,240 (6.2%) presentations due to MI with similar proportions in the control (6.2%) and intervention (6.1%) arms. Demographics for each site were consistent across arms (Tables S6 to S11).

**Figure 3.**
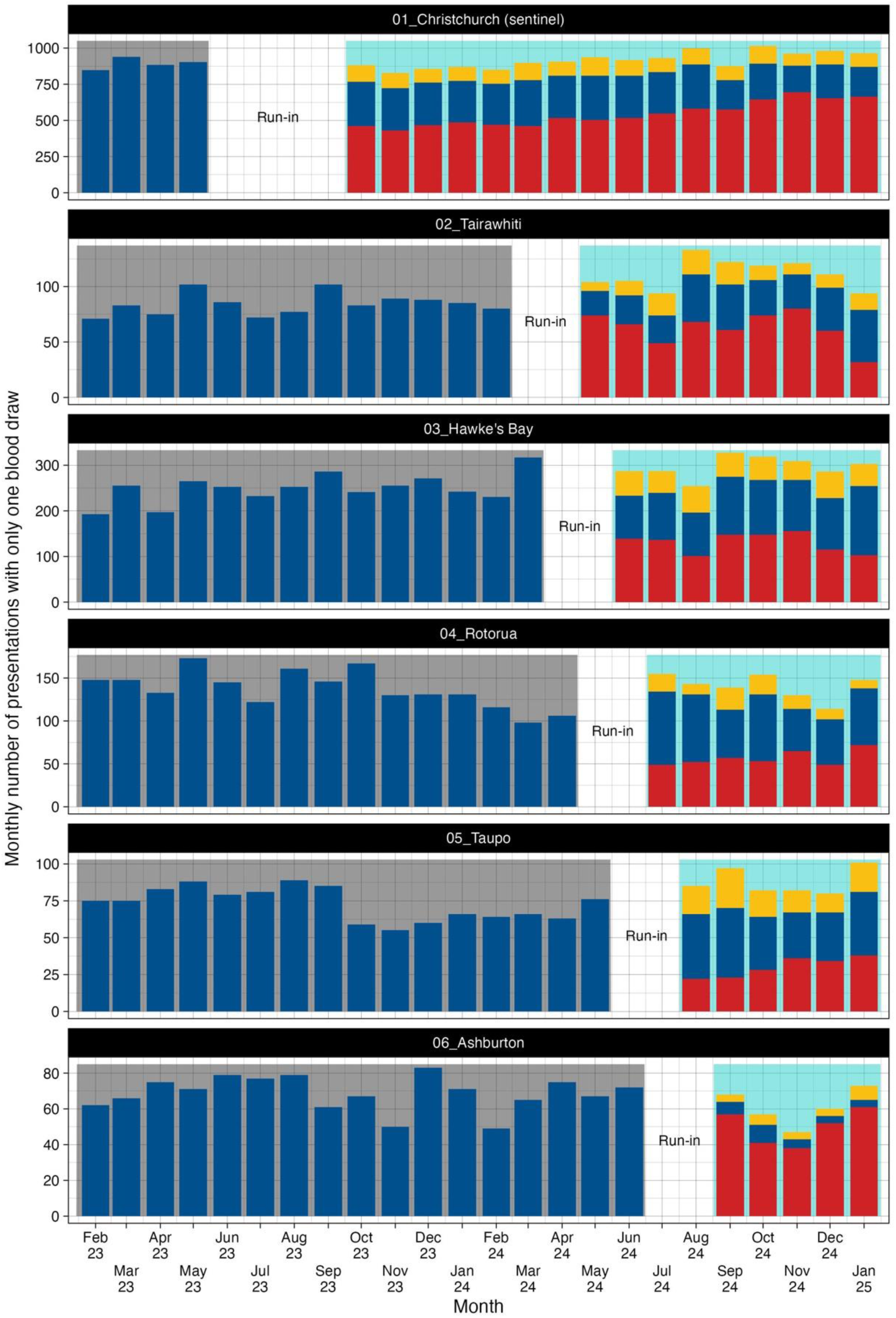
Central-laboratory and POC hs-cTn testing by site and trial phase: monthly numbers of presentations with only one blood draw. Blue columns are presentations with central-laboratory test only, Red with POC test only, and Yellow with both central-laboratory and POC. Grey background is the control arm, teal background the intervention arm. The gap is the run-in phase.

**Table 1.** Characteristics of all study participants at their first presentation.

| **Characteristics** | **Total**<br>**(N=44,747)** | **Control**<br>**(N=17,513)** | **Run-in**<br>**(N= 5,699)** | **Intervention**<br>**(N=21,535)** |
| --- | --- | --- | --- | --- |
| Age, mean ± standard deviation | 61 ± 19 | 61 ± 19 | 61 ± 19 | 60 ± 19 |
| Sex, Female | 22,168 (49.5%) | 8,648 (49.4%) | 2,863 (50.2%) | 10,657 (49.5%) |
| Ethnicity* |  |  |  |  |
| Māori | 8,501 (19.0%) | 4,357 (24.9%) | 842 (14.8%) | 3,302 (15.3%) |
| Pacific Peoples | 1,599 (3.6%) | 637 (3.6%) | 229 (4.0%) | 733 (3.4%) |
| Asian | 2,922 (6.5%) | 837 (4.8%) | 409 (7.2%) | 1,676 (7.8%) |
| Middle Eastern/Latin American/African | 480 (1.1%) | 134 (0.8%) | 65 (1.1%) | 281 (1.3%) |
| European | 31,049 (69.4%) | 11,499 (65.7%) | 4,129 (72.5%) | 15,421 (71.6%) |
| Other | 125 (0.3%) | 23 (0.1%) | 19 (0.3%) | 83 (0.4%) |
| Residual Categories | 71 (0.2%) | 23 (0.1%) | 6 (0.1%) | 39 (0.2%) |
| Triage Category** |  |  |  |  |
| 1 | 601 (1.3%) | 246 (1.4%) | 68 (1.2%) | 287 (1.3%) |
| 2 | 21,001 (46.9%) | 7,781 (44.4%) | 2,868 (50.3%) | 10,352 (48.1%) |
| 3 | 20,476 (45.8%) | 8,243 (47.1%) | 2,479 (43.5%) | 9,754 (45.3%) |
| 4 | 2,539 (5.7%) | 1,177 (6.7%) | 274 (4.8%) | 1,088 (5.1%) |
| 5 | 122 (0.3%) | 64 (0.4%) | 10 (0.2%) | 48 (0.2%) |
| Missing | 8 | 2 | 0 | 6 |

See Table S3 for more details and Table S4 for per presentation demographics.
\* The apparent disparity between study arms of ethnicity is because of when each site started the intervention and because the ethnic make-up varies considerably between hospitals. Ethnicity in between arms was balanced at each hospital (Supplemental Tables S5 to S10).
\*\* Australasian Triage Scale. Ranges from 1, immediately life-threatening, to 5, less urgent or administrative issues only.

The median (interquartile range) for the time from arrival to blood draw was 35 (20-65) minutes in the control arm and 33 (22-65) minutes in the intervention arm.

### PRIMARY and SECONDARY EFFECTIVENESS OUTCOMES

Length of stay was reduced by 13% (95%CI 9 to 16%; P<0.001) after implementation of the POC assay (Table S12, Fig.S4), corresponding with a 47-minute (95%CI: 33 to 61 minutes) reduction from the control arm’s mean length of stay of 376 minutes.

Length of stay was reduced by 7% to 15% in each of the secondary analyses (Fig.4, Table S13). The implementation increased odds of discharge within the 1, 2, 3, 4, 5, and 6-hour time windows (Fig.5, Table S14). Length of stay reduced by 8% (95%CI 1% to 14%) in sites excluding the sentinel site, 12% (95%CI 6% to 17%) in males and 11% (95%CI 6% to 16%) females when analyzed separately, and 14% (95%CI 7% to 21%) when the analysis was restricted to the youngest age group (Fig.4, Tables S15-S17). In sub-groups stratified by ethnicity, the reduction in length of stay was lower for Asian and Māori than for European (Fig.4, Table S18).

**Figure 4.**
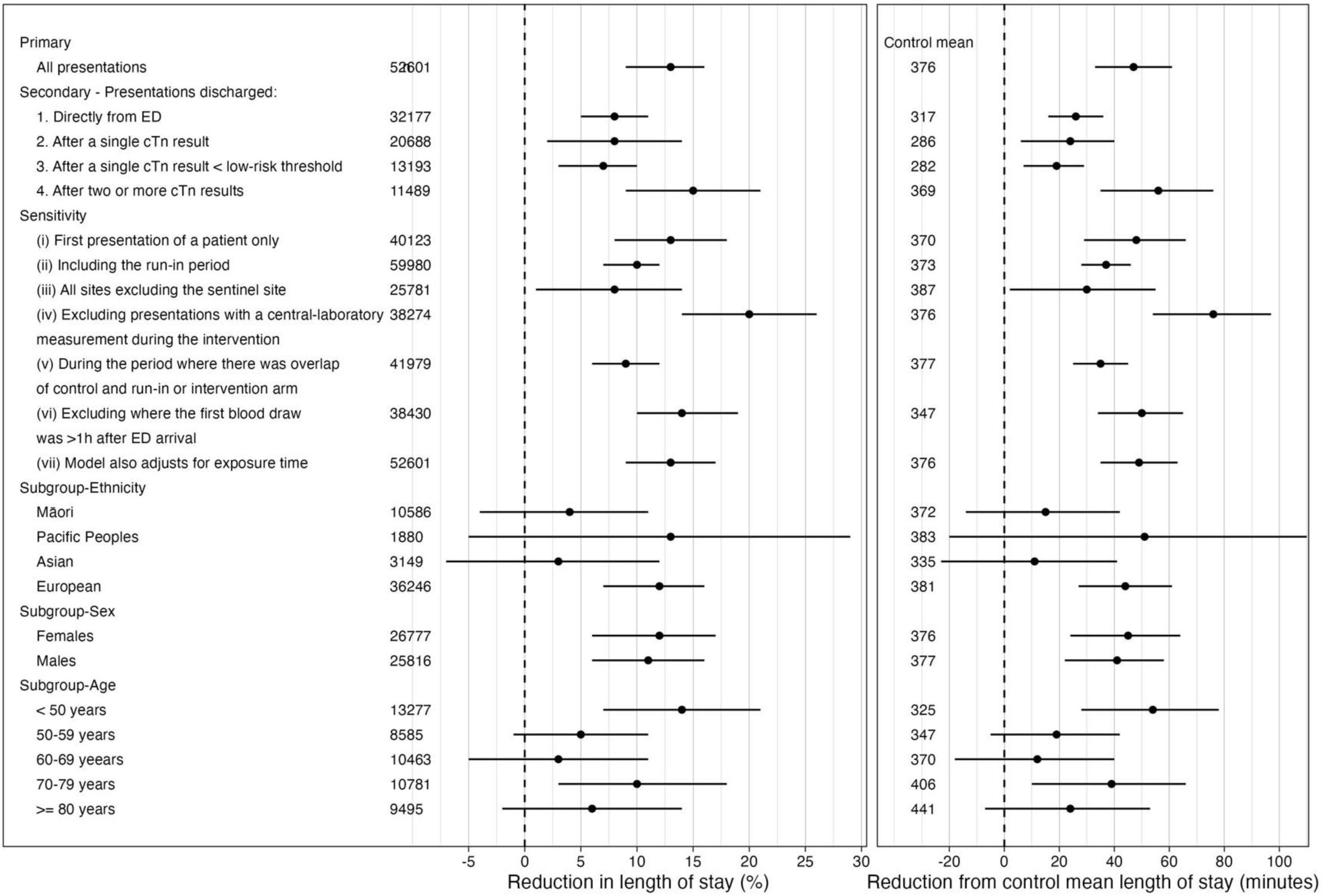
Reduction in ED length of stay. Primary effectiveness and key secondary effectiveness analyses, sensitivity analysis, and demographic sub-groups (see Supplemental Appendix for more details). Percentage reduction is the key outcome from the statistical model. The second panel is the reduction in control cohort mean length of stay in minutes. This is calculated by multiplying the percentage reduction by the control mean length of stay.

**Figure 5.**
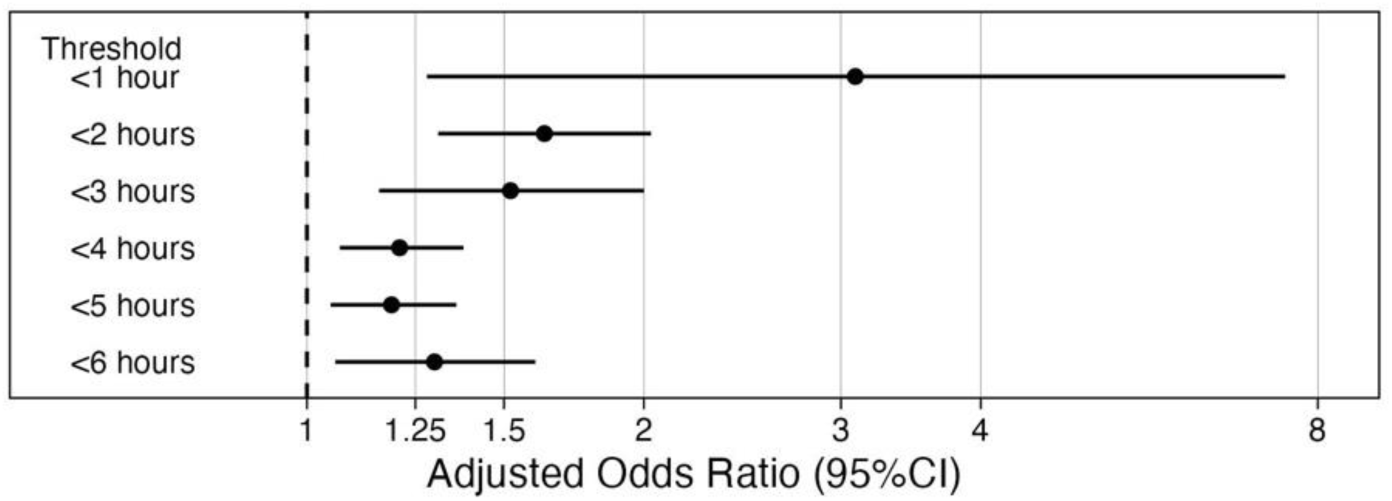
Change in odds for leaving the ED within 1-to-6-hour time windows.

### SAFETY OUTCOME

The rate of MI or cardiac death at 30 days for patients discharged from ED was similar in the control and intervention (50/12,906 [0.39%] versus 76/19,271 [0.39%], P=0.54) arms (Table 2). The rate of MI or all cause death at 30 days for patients discharged from ED was similar in the control and intervention (67/12,906 [0.52%] versus 110/19,271 [0.57%], P=0.86) arms.

**Table 2.** Safety Outcomes – 30-day MI or cardiac death following a presentation in which the patient was discharged from ED.

| **Outcome**<br>**(30-day)** | **Control**<br>**N(%)** | **Intervention**<br>**N(%)** | **Percentage points**<br>**difference (95%CI)** |
| --- | --- | --- | --- |
| Total presentations | 12,906 | 19,271 |  |
| MI | 43 (0.33%) | 67 (0.35%) | 0.06 (-0.12 to 0.15) |
| Cardiac death | 7 (0.05%) | 9 (0.05%) | -0.01 (-0.06 to 0.05) |
| Total events | 50 (0.39%) | 76 (0.39%) | 0.01 (-0.14 to 0.15) |

In the control arm the re-presentation to an ED rate was 18.4% and in the intervention arm it was 17.9% (p=0.10).

## DISCUSSION

This is the first multi-center randomized trial to evaluate the effectiveness and safety of a POC hs-cTnI assay in routine clinical care. Implementation of POC hs-cTnI reduced length of stay by 13%, a 47-minute reduction compared to using a central laboratory assay. The reduction in length of stay was safe with no increase in 30-day MI or cardiac death. Such a change is clinically meaningful and occurred despite a 23.4% increase in overall ED attendance. If applied across New Zealand (population 5.3 million), this reduction could reduce ED occupancy by ∼50,000 hours annually.

A reduction in length of stay was present both in patients discharged after a single troponin test and after serial testing. Three-quarters of presentations in the intervention arm were evaluated using the POC assay, with some variation in uptake by site. Further research is needed to understand differences in adoption.

The intervention performed equally well in males and females. An apparent difference in performance across ethnicities may, in part, relate to variation in POC use at sites with different Māori populations. Christchurch and Ashburton, which had the highest POC use, had the lowest Māori populations. Clinicians in New Zealand are cognizant that Māori and Pacific Peoples, like many indigenous populations, have higher rates of underlying coronary disease than non-Māori/non-Pacific Peoples.^22^ In three sites Māori or Pacific ethnicity is explicitly taken into account when assessing the risk of a cardiac event. This may have led to additional investigations and caution, prolonging length of stay. A decreased impact of ADPs on length of stay for Māori and Pacific Peoples was previously noted.^16^

### CONTEXT OF OTHER STUDIES

These findings suggest a much greater impact on length of stay than a recent single-center patient-level randomized controlled trial in Norway, which compared use of the VTLi hs-cTnI assay with a laboratory hs-cTnT assay finding a 6 minute (95%CI -4 to 17 minutes) reduction in median length of stay.^23^ However, there was already a short length of stay in the control arm; a median 3-hours compared to 5-hours in our study. In addition, to create a direct comparison with existing standards of care, there was no single-test rule-out process included because that was not the standard of care at that institution. Consequently, it was not possible to rule out patients after a single sample, and the potential benefit of this decision step may have been lost. Nevertheless, we observed in patients with serial samples a decrease in length of stay, a contrast to the Norwegian results, although the cohort of those receiving serial tests was not identical between studies. Other, observational studies have demonstrated diagnostic accuracy of hs-cTn POC assays and the potential for results to be available ∼40-70 minutes earlier than laboratory assays^24,25^ and a 0/2h pathways and single-sample rule-out thresholds have been derived and validated.^26,27^

Rapid change of physician practice patterns is difficult and potential benefits of a new diagnostic strategy can be obscured by variability in its use.^9,16,28^Accordingly, ICare-FASTER was designed with an *a priori* methodological decision to designate one site as a sentinel site, with an extended run-in period, and to analyze and refine the implementation strategy.^10,29^ First, we established that the VTLi assay had satisfactory analytical performance^14^ and determined a low-risk threshold suitable for use within ADPs.^15^ Despite this extensive preparation, we found that, as with many complex medical interventions, there were differences in uptake, as illustrated by variation in proportion of patients assessed using only POC testing during the intervention period. Future work to assess barriers to use of POC testing and their association with length of stay are needed.

The impact of interventions is influenced by the implementation strategy applied. We believe comprehensive pre-project stakeholder discovery, engagement and collaboration is essential. Specific attention to the following issues is required: (A) proactive communication to address concerns about perceived greater imprecision in POC assays than laboratory assays,^14,30,31^ (B) laboratory integration, (C) device IT connectivity, (D) logistics, device management and problem-solving, and (E) practical implementation issues such as accreditation, user education, and site-specific nuances of integration. Particular attention is needed regarding how clinicians interpret the POC and laboratory test results in combination with each other, as discordance between results is inevitable.^32,14^ The POC test invalid rate (8.6%) may have reduced the impact of the intervention by resulting in either a POC re-measurement or use of the laboratory troponin test. This may have delayed decision-making.

Prior to study initiation, concerns were raised that POC troponin testing would increase nursing workload by adding an extra task for nurses. We therefore undertook time-and-motion observations across 22 patient journeys involving patients presenting with chest pain, and measured the amount of time nurses spent performing tasks directly related to the care of each individual patient while awaiting laboratory results and physician decision-making. We found the time taken to use the VTLi (∼3 minutes) is much less than the time taken on tasks while waiting for laboratory-based decision-making (∼21 minutes on average), resulting in a net reduction of ∼18 minutes nurse time with a patient.

Performance gains in individual hospitals may vary depending on how quickly current processes deliver blood samples to a laboratory and how quickly those samples are analyzed and reported. In our hospitals, which prioritize troponin requests, this is typically one-hour and results are usually not available when initially looked for by clinicians. In our preliminary study in one hospital the median time from blood draw to posting results was 71 minutes for the laboratory test and 11 minutes for the POC test.^29^ It is possible that changes to laboratory procedures may narrow that gap and so, perhaps, reduce the effectiveness of the intervention.

### LIMITATIONS

During the study ED presentations increased substantially, bringing increased pressure on already busy EDs. To address this, the model adjusted for the time-from-arrival-to-first-blood-draw as a proxy for ED workload, and allowed the secular time variable to vary with site and be modelled by non-linear splines.^33–36^ Also, we adhered to a statistical analysis plan and conducted multiple pre-specified sensitivity analyses to support the robustness of the primary findings. This was only a single country study. However, New Zealand has a similar model of Emergency Medicine specialist-led ED care delivery to other major developed nations, including Australia, Canada, the United Kingdom and the USA.

This study was conducted using only one hs-cTn POC test. Nevertheless, we would expect other hs-cTn POC tests with comparable diagnostic accuracy and analytical sensitivity to reduce length of stay even if they had slightly longer analytical turnaround time, as long as results are available when looked for by the clinician to enable decision-making.. These findings may vary with ADP and so may not be generalizable to EDs that rely on other pathways such as the European Society of Cardiology algorithms. Finally, no deliberate changes to work-flow were made. Such changes may produce additional time-saving and other benefits.

Normally patients with clear ST-segment elevation of the electrocardiogram do not receive a troponin test before transfer to coronary angiography. However, sometimes a troponin test is made in these patients and so we did not exclude them. This may have reduced the effectiveness of the intervention as POC testing would not alter the length of stay in this small subgroup of patients.

## CONCLUSION

Implementation of POC whole-blood testing using the Atellica VTLi hs-cTnI safely reduced ED length of stay when integrated into ADPs for the assessment of possible MI.

## Data Availability

Data will not be shared or will be available upon request.

## ACKNOWLEDGEMENTS

We thank Alieke Dierckx for her unceasing administrative support throughout the literal hundreds of meetings; Carol Limber for management support; Rachael Johnson for the professional project management; Dr Richard Parker for discussions concerning statistical modelling; NZ Health National Collections Data Service for data provision; Ben Briggs, Mary Crowe, and Matthew Dean for their technical support to integrate the POC devices with the clinical record; Minh Phan, Felicity Taylor, Chris Finaly from Canterbury Health Laboratories for analyser support, Dr Campbell Heron, Dr Stephen Du Toit, Dr Tim Sutton, Professors Kristin Aakre, Rob Christenson, Paul Collinson and Dr Leo Lam for advice on pre-study verification of POC devices.; Dr R Alex Stothart and Otis Williams for outcome adjudication in the pre-study POC threshold determination phase; and Professor Peter George for support with the initial grant application. NLM is supported by the British Heart Foundation through a Chair Award (CH/F/21/90010), a Programme Grant (RG/20/10/34966) and a Research Excellence Award (RE/24/130012).

